# Intraoperative nerve imaging with sodium fluorescein

**DOI:** 10.1101/2025.02.08.25321923

**Authors:** Roy K. Park, Bernardo A. Arús, Jennifer Y. Lee, Merle M. Weitzenberg, Melissa C. Lee, Mark S. Nyaeme, Julia Barthel, Giuseppe Balsamo, Fred M. Baik, Katie Speirs, Berit Blume, Metar Heller-Algazi, Andriy Chmyrov, Oliver Plettenburg, Uchechukwu C. Megwalu, Jürgen Weitz, Marius Distler, Oliver T. Bruns, Tulio A. Valdez

**Affiliations:** Department of Otolaryngology, Head and Neck Surgery, Stanford University, Palo Alto, CA, USA; National Center for Tumor Diseases (NCT/UCC), Dresden, Germany; German Cancer Research Center (DKFZ), Heidelberg, Germany; Medizinische Fakultät and University Hospital Carl Gustav Carus, Technische Universität Dresden, Dresden, Germany; Helmholtz Zentrum Dresden-Rossendorf (HZDR), Dresden, Germany; Helmholtz Pioneer Campus, Helmholtz Zentrum München, Neuherberg, Germany; Institute of Medicinal Chemistry, Molecular Targets and Therapeutics Center, Helmholtz Zentrum München, Neuherberg, Germany; Institute of Organic Chemistry, Center of Biomolecular Research (BMWZ), Leibniz University Hannover, Hannover, Germany; Department of Anesthesiology, Perioperative and Pain Medicine, Stanford University, Palo Alto, CA, USA; Department of Visceral, Thoracic and Vascular Surgery, University Hospital and Faculty of Medicine Carl Gustav Carus, Technische Universität Dresden, Dresden, Germany; Laboratory of Nano- and Quantum Engineering (LNQE), Leibniz University Hannover; Hanover, Germany; Institute of Lung Health (ILH); Gießen, Germany; German Center for Diabetes Research (DZD); Neuherberg, Germany

## Abstract

Nerve damage during surgery is a common and serious complication, often leading to chronic pain, functional impairments, and diminished quality of life. However, existing methods for intraoperative nerve identification remain insufficient, especially for detecting small or hidden nerve branches. Here we present a new application of a clinically approved fluorescent agent, sodium fluorescein, to enhance nerve visualization during surgery. Utilizing both clinical and customized imaging systems, fluorescein remarkably improved nerve contrast, revealing structures undetectable with white light, including small branches embedded within tissues. With its established safety profile, low cost, and immediate clinical applicability, sodium fluorescein offers the potential to revolutionize surgical practice by minimizing nerve injuries and improving patient outcomes. Clinical Trial Registration: NCT06054178.

**Graphical abstract:** 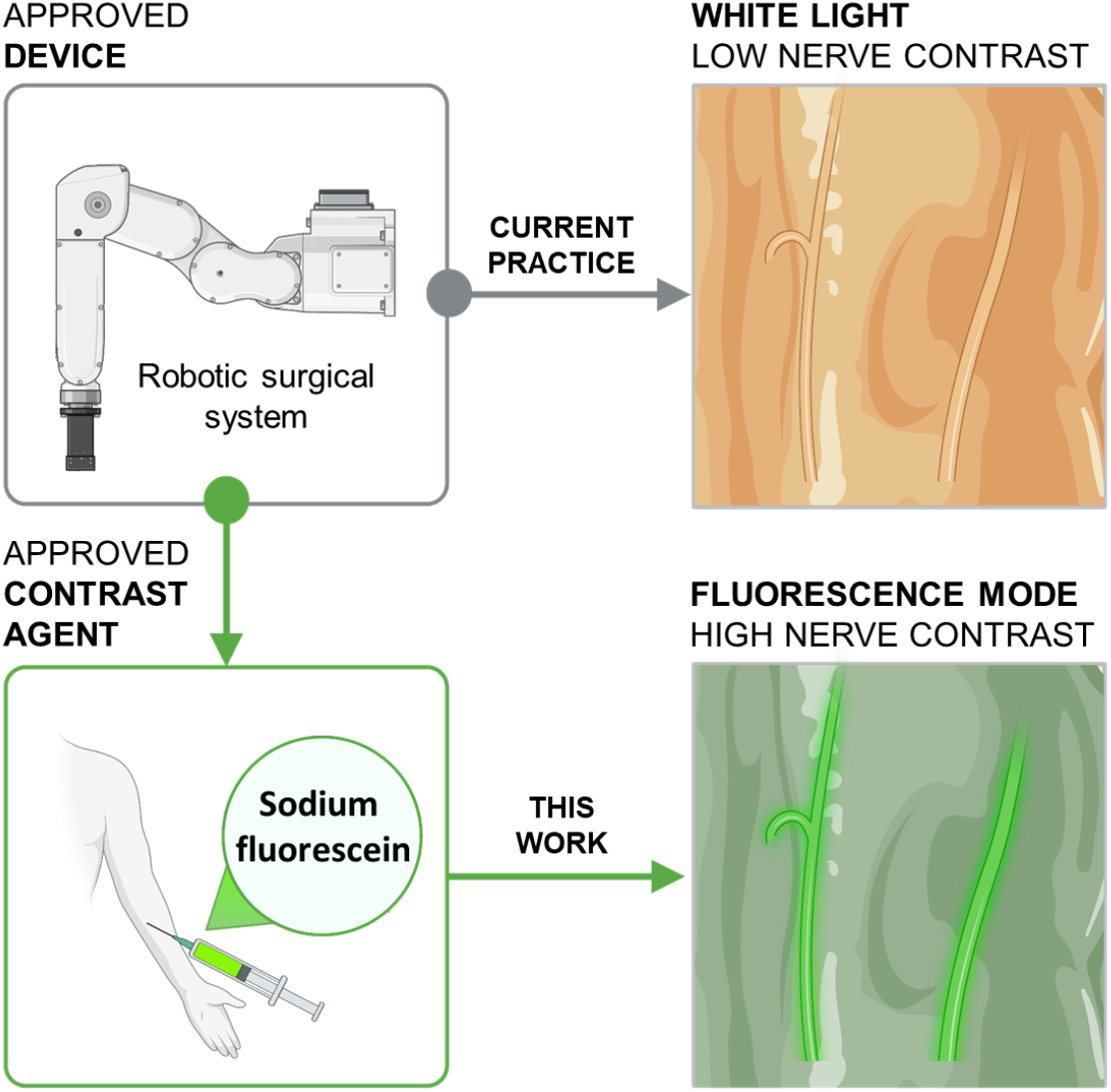

---

Nerve damage in surgical procedures can severely impact patient quality of life, causing chronic pain, motor and sensory deficits, or permanent loss of function^1,2^. For example, surgeries in the region of the pelvis minor – like colorectal cancer surgeries and gynecological procedures - pose a high risk to pelvic nerves, resulting in sexual dysfunction and urinary or fecal incontinence^3-5^. Moreover, head and neck surgeries risk damage to cranial nerves, leading to swallowing difficulties, voice changes, or even facial paralysis^2,6^.

Despite advancements in surgical techniques, intraoperative nerve visualization and protection remains a critically unmet need^1,2^. Current methods for nerve identification rely on visual inspection and knowledge of anatomical landmarks, which can be challenging when nerves are obscured by surrounding tissues or pathological changes. With minimally invasive approaches in prostate or pelvic surgery, nerves may never be directly visualized and are especially susceptible to blunt or thermal injury when solely relying on anatomical landmarks^7^. Intraoperative monitoring tools such as electromyographic nerve monitoring only provide feedback after nerve contact or damage has occurred and does not allow for identification of sensory nerves^8^. While efforts to develop novel nerve targeting agents are underway, no agents have yet received regulatory approval, leaving surgeons to rely on visual inspection^9-11^.

In this study, we report a novel application of sodium fluorescein, a clinically approved fluorescent contrast agent, to improve intraoperative nerve visualization. Sodium fluorescein is inexpensive, presents a well-established safety profile, and has been widely used in ophthalmology^12-14^. By employing clinical imaging systems and administering sodium fluorescein at standard doses, we demonstrate the ability to visualize nerves with superior contrast compared to conventional visible-light inspection. Importantly, this approach allows for the identification of nerves even when hidden within fatty or fibrous tissue. Since both sodium fluorescein and imaging devices are already clinically approved, this approach enables widespread and prompt adoption, offering immediate potential to reduce nerve damage and change the current practice of surgery.

To evaluate the ability of sodium fluorescein for nerve visualization, we conducted a feasibility clinical trial on the visualization of cranial nerves in patients undergoing salivary gland tumor removal (**Figure 1a**). Six patients were administered 1 mg/kg of intravenous sodium fluorescein at anesthesia induction and underwent imaging sessions between 2 and 3.5 hours after administration **(Table S1)**. Using a custom imaging setup (**Figure S1**) optimized to enhance fluorescence signal while minimizing reflected light, we successfully visualized the facial nerve (CN VII) with greater contrast compared to visible light alone (**Figures 1, S2)**. Weber contrast ratios to quantify facial nerve fluorescence compared to background tissues (mean ± SD) was 2.5 ± 1.0. Notably, we were able to highlight small branches of the facial nerve around 1 to 2 mm in size, which were difficult to detect with visible light alone (**Figures 1b-1d**). All detected facial nerve branches were further confirmed intraoperatively via electromyography at 0.5 mA stimulation.

**Figure 1.**
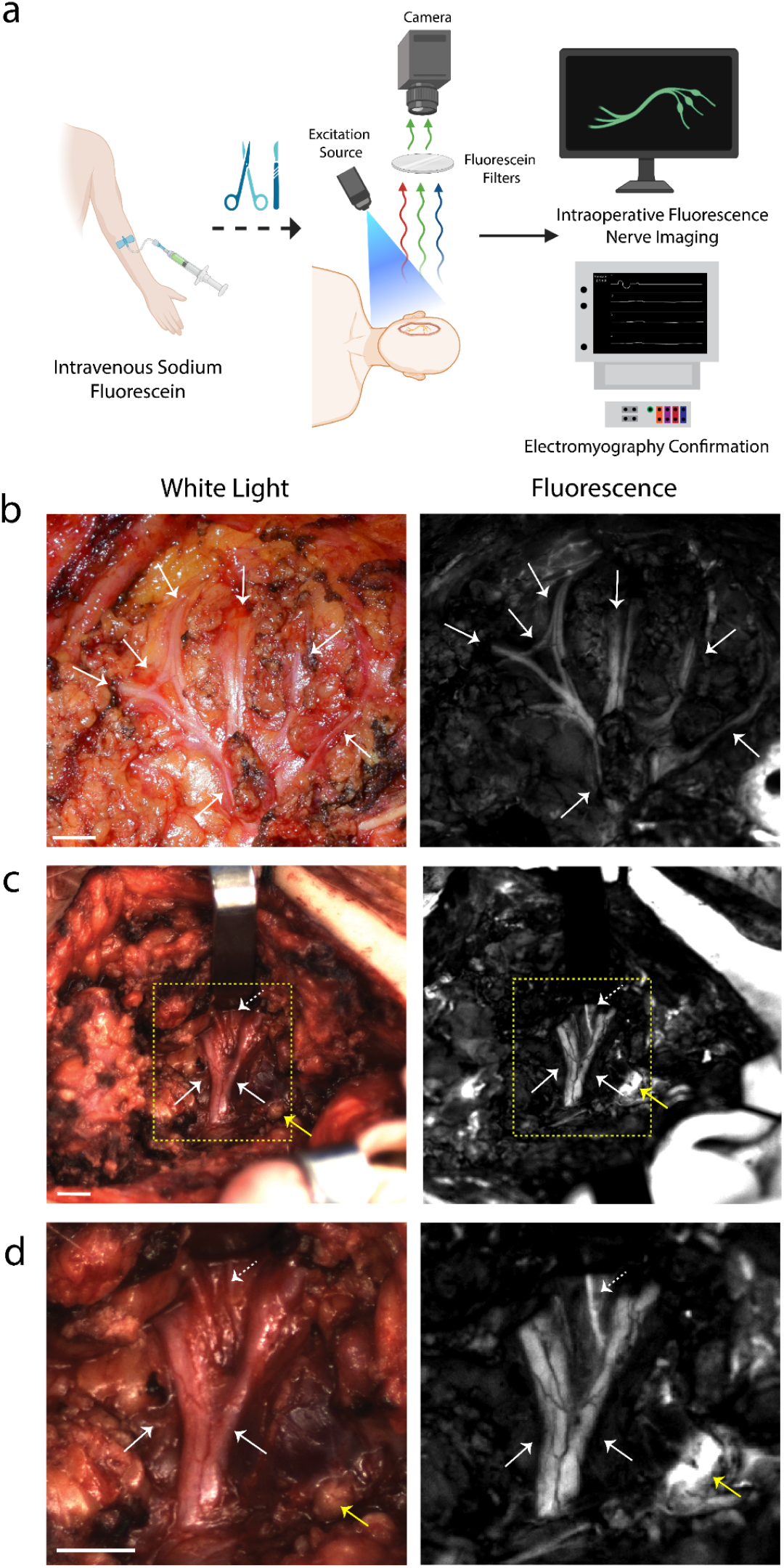
Fluorescence nerve imaging with sodium fluorescein during parotidectomy. a) Schematic representation of nerve imaging clinical trial. Patients were administered sodium fluorescein at anesthesia induction and subsequently imaged with a custom imaging set up once nerves are exposed intraoperatively. Image created on Biorender.com. b-d) Visible light and corresponding fluorescence images of exposed CN VII (facial nerve). White solid arrows represent nerves, while the dotted white arrows point to nerve branches not discernable by visible light alone. Yellow arrow corresponds to lymph node. The corresponding patient information including imaging timepoints after sodium fluorescein administration is available on Table ST1. The displayed contrast was optimized for each fluorescence image and bands with corresponding intensity are displayed in Fig. S2. Scale bar 0.5 cm.

To assess the viability of clinically approved commercial imaging systems to visualize nerves highlighted with sodium fluorescein, we conducted a comparative experiment using the Zeiss Kinevo microscope equipped with the Yellow 560 filter, a commonly utilized system for fluorescein imaging. The Kinevo microscope was able to detect strong nerve fluorescence signals compared to surrounding tissues (**Figure S3**). However, it failed to visualize finer nerve branches due to masking light reflections which are stronger than the fluorescence signal at this dose of 1 mg/kg (**Figure S3)**.

To further extend the application of sodium fluorescein imaging to robotic-assisted surgeries, we employed the Da Vinci Xi surgical system, focusing on nerve visualization of the superior hypogastric plexus in a patient undergoing a robotic low anterior resection. Using the 30° endoscope in standard mode, we set the lowest white light and highest Firefly fluorescence intensities to optimize imaging conditions. The patient was intravenously injected with a dose of 5 mg/kg sodium fluorescein at anesthesia induction to ensure adequate fluorescence signal strength. This setup successfully increased the contrast of pelvic nerves at approximately 1.5 h hours after injection, revealing structures otherwise not visible under white light (**Figures 2, S4, Supplementary Video S1**). Notably, photobleaching from fluorescein was not detected during the surgery under at least 30 minutes of continuous illumination. Additionally, experiments comparing the Da Vinci Xi to a customized endoscope system highlight that slight optimization of the light source and detection filters can further improve nerve detection sensitivity in minimally invasive settings (**Figures S5, S6)**.

**Figure 2.**
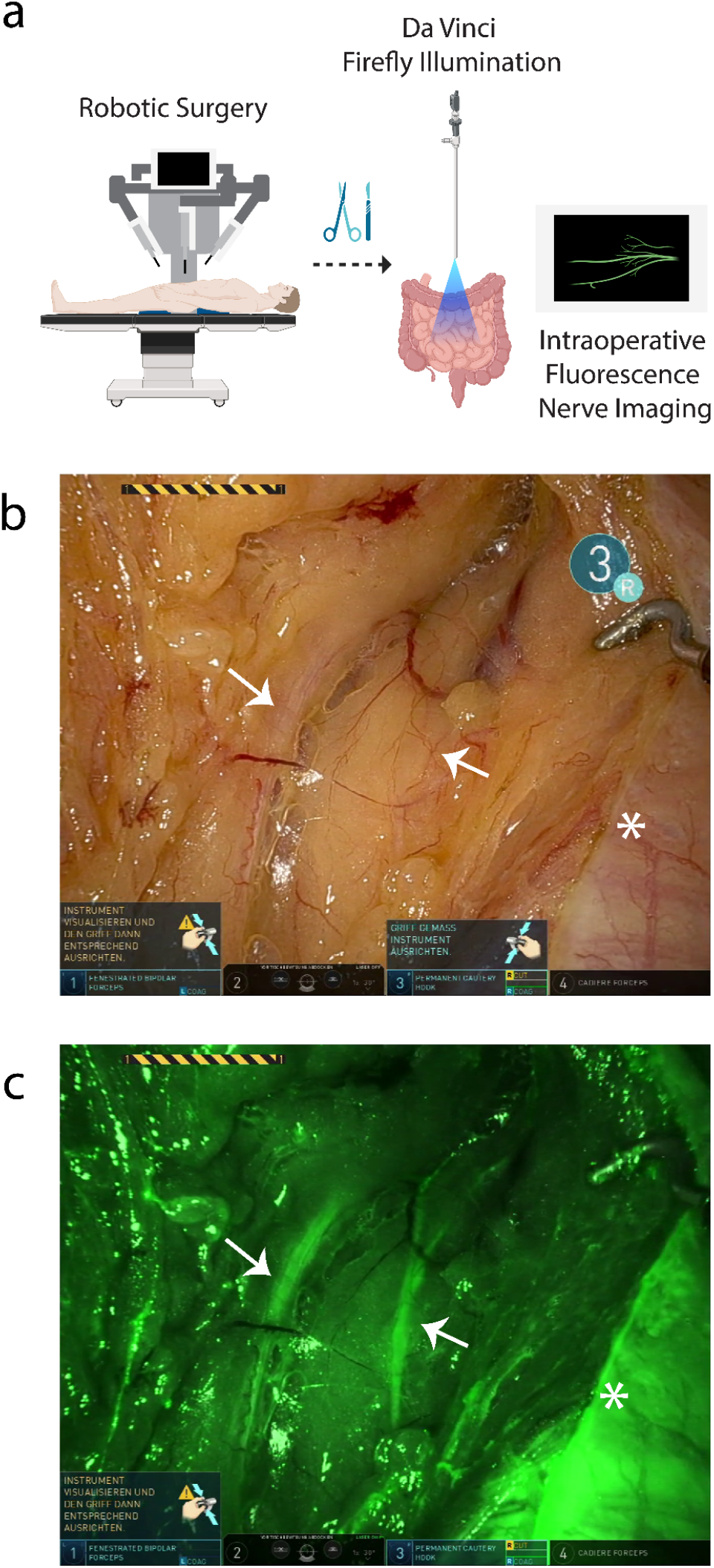
Nerve imaging with Da Vinci Firefly endoscope after fluorescein administration in pelvic surgery. a) Schematic representation of fluorescence nerve imaging during robotic surgery after administration of sodium fluorescein. Image created on Biorender.com. b) White light images with Da Vinci endoscope during pelvic surgery c) Corresponding fluorescence images with the Da Vinci Firefly fluorescence endoscope. Settings were set at maximum Firefly illumination and lowest white light settings to allow for visualization of nerves of the superior hypogastric plexus otherwise not visible under white light alone. White arrow corresponds to nerves of the superior hypogastric plexus. White star is margin of opened peritoneum in the small basin.

To determine feasibility and potential for translation prior to clinical trial enrollment, we also tested our hypothesis to use sodium fluorescein to identify nerves in murine pre-clinical models. Fluorescence from sodium fluorescein administration improved nerve contrast compared to visible light alone **(Figure S7)**. The results from the pan-body survey of nerve fluorescence suggest that this nerve identification technique can be applied to any anatomical region or surgery type.

In both our pre-clinical and clinical data, we noted other structures that also provided fluorescence intraoperatively after the administration of fluorescein. Artifacts from tissue electrocauterization yielded a fluorescence intensity similar to that of nerves (**Figures 2a, S3, S4, S8**). This cautery artifact, however, was easily distinguished from nerves under white light given the darkened appearance of cautery. Similarly, overlying fascia also showed fluorescence but could be distinguished from nerves by visual inspection and shape (**Figures 2, S4**). Lymph nodes also provided fluorescence but were easily distinguished from nerves given their differences in structure (**Figure 1d)**. In robotic surgery, the ureters were expectantly fluorescent given the renal excretion clearance pattern of fluorescein (**Figure S4b**), which we consider an additional benefit as this signal can be used to avoid ureteral injury.

With current clinical practice, fluorescein via intravenous administration has traditionally been used for ophthalmic angiography but has also recently gained traction in peripheral nerve schwannoma resection and nerve biopsies^12,13,15,16^. However, in these applications, the use of sodium fluorescein and the imaging conditions were not optimized for the visualization of healthy nerves^17,18^. As a result, non-pathological nerves displayed only mild fluorescence, making them difficult to distinguish from surrounding tissue, particularly smaller branches that were obscured by white light reflections.

Our study demonstrates that high-contrast imaging of fluorescein-labeled nerves is possible intraoperatively with currently approved clinical imaging systems, particularly in robotic surgery using the Da Vinci Firefly system. Effective nerve visualization was achieved after intravenous doses of 1 and 5 mg/kg, consistent with current clinical applications in ophthalmology and tumor imaging^13,15,18^. Logistically, sodium fluorescein can be given at anesthesia induction without disrupting the surgical workflow, reducing further burden on the patient. Moreover, wide availability, low cost, and compatibility with existing clinical imaging systems make fluorescein especially valuable for resource-limited settings, where cost-effective solutions are essential^14^.

Notably, this work shows that custom imaging systems optimized to minimize reflections enabled the visualization of not only peripheral nerves but also small branches that were undetectable under visible light. These findings suggest that minor adjustments in commercial imaging platforms, for instance by optimizing fluorescence filters to remove non-fluorescent light such as reflections and excitation light bleed through, have the potential to remarkably improve nerve visualization in open and robotic-assisted procedures.

The ability of sodium fluorescein to highlight small nerve branches and those concealed within fatty tissues have immediate clinical relevance. Surgeries with high rates of nerve injury or with a high-cost outcome caused by nerve damage, such as gynecological procedures and rectal resections, stand to benefit the most from this technique. Our findings pave the way for widespread clinical adoption in select clinical cases, with the potential to transform the current landscape of nerve preservation across a broad range of surgical specialties. Further randomized controlled clinical trials will assess nerve preservation outcomes using fluorescence-guided surgery after the administration of sodium fluorescein.

Considering the unmet need to enhance intraoperative nerve visualization, several new nerve-targeting contrast agents have been under development in the past decades^9-11,19^. These next-generation dyes are designed to achieve high specificity and strong fluorescence contrast to surrounding tissue, potentially enabling even more precise nerve identification than fluorescein. However, given the immediate translational potential of fluorescein, it stands as an important benchmark for the development of novel dyes and may offer a practical solution until improved agents become clinically available.

In conclusion, our study demonstrates that sodium fluorescein, a readily accessible dye, enhances intraoperative nerve visualization. No approved methods currently exist to identify nerves via fluorescence intraoperatively with high contrast. Fluorescein may provide surgeons with a real-time tool to reduce nerve injury risk with minimal disruption to standard clinical workflows. While nerve injury remains one of the most dreaded complications in surgery, the immediate translational nature of this study has the potential to significantly improve patient outcomes and quality of life across a wide range of surgical specialties.

## Material and methods

### Custom Clinical Fluorescence Imaging Set Up

This imaging setup (**Figure S1**) utilized a Teledyne FLIR Blackfly S USB3 12 MP camera (BFS-U3-120S6M-C) controlled via MicroManager. The camera operated without cooling, gain, or gamma adjustment (gamma set to 1) with binning set at 2. A 35-mm camera lens (f/1.4, Thorlabs, catalog no. MVL35M1) was used. The emission pathway was designed for optimal transmission in the range between 500 and 550 nm, achieved using three main filters: a long-pass (LP) 500 nm filter (Thorlabs, catalog no. FELH0500), colored glass LP filter 495 nm (Thorlabs, catalog no. FGL495M), and a short-pass (SP) 550 nm filter (Thorlabs, catalog no. FESH0550). Illumination was delivered by two 470 nm LEDs (M470L5, Thorlabs) with a maximum power density of 12 mW/cm^2^ across the field of view. Each LED was equipped with an ACL2520U lens, positioned slightly out of focus to create even illumination. To clean-up the excitation wavelength range, a band-pass (BP) filter 469-35 (Thorlabs, catalog no. MF469-35) was used. Images were saved as 12 bit (0 to 4095). Visible images were taken with an Allied Vision Alvium 12 MP color camera (1800 U-1240c) with a 25 mm camera lens (f1.4, Thorlabs, catalog no. MVL25M23) or Nikon DSLR 3300 camera with a 35 mm lens.

### Head and Neck Nerve Imaging Clinical Trial

The clinical trial was approved by the Stanford Institutional Review Board (IRB 71857) and the study was registered on ClinicalTrials.gov (ClinicalTrials.gov identifier: NCT06054178). Eligible patients undergoing head and neck surgery older than 18 years provided written informed consent. Patients with history of fluorescein allergy or history of kidney disease were not eligible to participate in the study. No adverse effects related to dye administration were observed for enrolled patients in the study.

Upon induction of anesthesia, patients were administered an intravenous dose of 1 mg/kg sodium fluorescein (Fluorescite Injection 10%). All medications were obtained from the Stanford Investigational Drug Service (IDS) pharmacy. No long-term paralytics were administered to allow for nerve monitoring, and sedation levels were maintained with total intravenous anesthesia (TIVA). Facial nerve function was monitored intraoperatively via electromyography (EMG) with the NIM nerve vital monitoring system (Medtronic). Following dissection and identification of the main trunk of the facial nerve, nerve imaging was performed with our custom imaging system at an exposure time of 50 to 100 ms. Nerve branches identified on the custom imaging system that were not visible with white light were further verified with EMG stimulation at 0.5 mA. To prevent any light contamination, all computer screens and overhead lights were turned off during fluorescence imaging.

As a benchmark, the Yellow 560 fluorescence system on the Zeiss Kinevo was also compared to our custom system intraoperatively. Images were acquired at the auto shutter setting.

### Robotic Colorectal Surgery

Informed written consent with explicit consent for publication of data was obtained from a single patient undergoing colorectal robotic surgery with the Da Vinci Xi at the University Hospital Carl Gustav Carus of the Technical University Dresden. Following induction of anesthesia, the patient was administered an intravenous dose of 5 mg/kg sodium fluorescein (Fluorescite Injection 10%). Fluorescence imaging was performed with the Da Vinci Xi Firefly Fluorescence system on Firefly Standard mode with settings on lowest white light and highest Firefly fluorescence intensities. No adverse effects from fluorescein administration were observed.

### Animal Imaging

Murine protocols were approved by the Stanford Administrative Panel on Laboratory Animal Care. Further murine protocols were approved by the government of Upper Bavaria, Germany. All procedures complied with ARRIVE (Animal Research Reporting of In Vivo Experiments) guidelines.

### Murine Fluorescein Studies

6 to 8 week old balb/c mice (Jackson Laboratory) were anesthetized with 2% v/v isoflurane and intravenously administered 15 nmol/gram sodium fluorescein (Sigma-Aldrich) via retro-orbital injection. 5 hours following administration of sodium fluorescein (n = 5), mice were euthanized with carbon dioxide in a chamber provided by the Stanford animal research facility. Following this, pan-body surveys of nerves were performed with both white light and fluorescence imaging. All imaging for the murine experiments were performed using the custom set up (Imaging Setup A). All images were acquired at 100 ms.

### Imaging Setup A (Murine Studies)

For murine studies, the same optical set up as the custom clinical intraoperative imaging device was used. All visible light images were taken with the Allied Vision Alvium 12 MP color camera (1800 U-1240c) with a 25 mm camera lens (f1.4, Thorlabs, catalog no. MVL25M23) on Vimba Viewer X (Allied Vision). For fluorescence imaging, the illumination was delivered with a maximum power density of 25 mW/cm^2^ across the field of view.

### Imaging Setup B (Custom Endoscope)

This custom fluorescein endoscope consisted of a Panoview Ultra endoscope with a 10-mm diameter and 30° head angle (Richard Wolf, catalog no. 8934462), connected to an Alvium 1800 U-508m 5.1mp monochrome camera (Allied Vision, catalog no. 14662) using a RIWO-Zoom Adapter (Richard Wolf, catalog no. 85261504). The camera was controlled via MicroManager (v. 2.0.3) and operated without cooling or gamma adjustment, at an exposure time of 30 ms and with gain set to 48 dB. Illumination from a 470-nm LED (Thorlabs, catalog no. M470L5, 810mW) was collimated, filtered through a 469-nm band pass filter with a 35-nm band width (Thorlabs, MF469-35), and focused into a liquid light guide (Richard Wolf, catalog no. 806550231) to the endoscope, with a maximum power density between 2.5 and 8 mW/cm^2^, depending on the field of view. Emission light was filtered with a 520nm band pass filter with a 36-nm band width (Edmund Optics, catalog no. 67-016) attached to the camera adapter. Care was taken to ensure no transmission overlap between the excitation and emission filters, to avoid reflections induced by the illumination source. Images were saved as 8 bit (0 to 255).

### *In Vitro* Sensitivity Tests with Custom Endoscope and Da Vinci Firefly

*In vitro* sensitivity tests were conducted with 10 µL capillary tubes (Hirschmann, catalog no. 9000110) filled with sodium fluorescein at concentrations ranging from 0.5 to 20 µg/ml, diluted in saline. The capillaries were positioned atop fresh excised porcine rib tissue, obtained from a local butcher. Comparative imaging was conducted using the custom endoscope (imaging setup B) and the Da Vinci Firefly Xi system with its integrated endoscope.

The Da Vinci Xi imaging system was evaluated under two configurations: the optimized setting (maximum Firefly illumination with minimal white light) and the default setting (50% white light and 50% Firefly illumination). To assess the impact of a more specific band-pass filter detection, images were acquired with the custom endoscope paired with Firefly illumination. Subsequent experiments were conducted using the custom endoscope’s illumination system to validate the combined effect of optimized illumination clean-up and band-pass filter imaging.

To further compare the ability of both imaging systems to detect fluorescein buried within tissue, a capillary containing 20 µg/mL sodium fluorescein was embedded approximately 1 mm below porcine tissue. Imaging was performed using the optimized configuration of the Da Vinci Firefly Xi system and the custom endoscope with custom illumination.

### Fluorescence Spectroscopy of Sodium Fluorescein

Sodium fluorescein was diluted to a concentration of 50 µg/mL. Using a TECAN Spark plate reader, excitation spectra was measured from 400 to 520 nm using a monochromator emission wavelength of 560 nm with a bandpass of 10 nm. Emission spectra was measured from 480 nm to 640 nm at an excitation of 440 nm with a bandpass of 5 nm. Samples were measured 5 times, normalized, and averaged.

### Data Processing

Images were post-processed utilizing Python 3 and Fiji distribution of ImageJ. For all images, 5 or 10 dark frames were acquired for each exposure time and averaged. These averaged dark frames were then used to subtract the dark current from each experimental image. Images were then further corrected for exposure time to yield units in counts/s. The displayed linear contrast on each image was optimized and adjusted in Fiji. All images were saved as BMP files on Fiji, then cropped and rearranged as figures on Adobe Illustrator.

### Image Analysis

Weber values for the fluorescence nerve images were calculated to define contrast with the following:

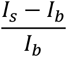

where I_s_ represents the specified region of interest while I_b_ represents the region of background.

## Acknowledgments

We would like to thank the generosity of patients for enrolling in the nerve imaging clinical trial. We would also like to thank Kalina Bamberg for support in the *in vitro* sensitivity experiment, Iuliia Mukha for support in the graphical abstract, as well as the Department of Neurosurgery at Stanford for use of the Zeiss Kinevo microscope and the Department of Anesthesiology at Stanford and at the University Hospital Carl Gustav Carus of the Technical University Dresden for assistance with sodium fluorescein administration.

## Contributions

OTB and TAV conceived the study. RKP and TAV wrote the IRB and registered the study on ClinicalTrials.gov. JYL, FMB, UCM, and RKP recruited patients in the Stanford clinical trial, and RKP obtained written informed consent for all patients. KS helped coordinate anesthesiology partners for intraoperative intravenous fluorescein injection. JB, MD, and JW provided the data from the Germany robotic surgery case. RKP and BAA built the custom intraoperative set up. RKP, JYL, MCL, MSN, FMB, and UCM performed the intraoperative imaging experiments. RKP, BAA, MMW, and MCL performed the murine experiments. MH and AC built the custom endoscope. BB and OP contributed to the initial stages of the project. RKP and BAA performed imaging processing and analysis. OTB and TAV supervised all experiments and guided the project. RKP, BAA, OTB, and TAV wrote the manuscript with input from all authors. All authors agreed to the publication of the manuscript.

## Data Availability

As generated data contains potentially identifiable patient information, raw data is not available due to patient confidentiality rules but can be considered with reasonable request to the corresponding authors.

