## Supplementary Material for "Intraoperative nerve imaging with sodium fluorescein"

**Contents:**

**Table S1** Demographics of patients enrolled in Stanford Clinical Trial (NCT06054178

**Figure S1** Custom intraoperative fluorescence imaging system

**Figure S2** Fluorescence nerve imaging of all patients enrolled in the Stanford clinical trial

**Figure S3** Fluorescence nerve imaging with Zeiss Kinevo Yellow 560 system

**Figure S4** Nerve imaging with Da Vinci Firefly endoscope

**Figure S5** Sensitivity comparison of the Da Vinci Firefly endoscope at default and optimized setting

**Figure S6** Comparison of Da Vinci Firefly endoscope and custom endoscope for embedded sodium fluorescein capillary tube under 1 mm of tissue

**Figure S7.** Pan-body survey of nerves in murine models after administration of sodium fluorescein

**Figure S8.** Electrocautery artifacts highlighted with sodium fluorescein imaging

**Supplementary Video S1** Representative section of the robotic low anterior resection with nerves of the superior hypogastric plexus highlighted with sodium fluorescein

**Supplementary Table 1** Demographics of patients enrolled in Stanford Clinical Trial (NCT06054178)

| Patient | Initial Diagnosis | Surgery | Nerve Imaged | Correlative Figure Images | Approximate Imaging Time Point after Dye Administration (min) | Final Pathology |
|---|---|---|---|---|---|---|
| Patient 1 | Bland oncocytic proliferation | Parotidectomy | CN VII | S2a | 190, 215 | Salivary oncocytosis |
| Patient 2 | Pleomorphic Adenoma | Parotidectomy | CN VII | 1c/1d, S2b | 140, 165 | Pleomorphic Adenoma |
| Patient 3 | Pleomorphic Adenoma | Parotidectomy | CN VII | S2c | 150 | Intracapsular epithelial-myoepithelial carcinoma (ex-pleomorphic adenoma) |
| Patient 4 | Pleomorphic Adenoma | Parotidectomy | CN VII | S2d | 175 | Pleomorphic Adenoma |
| Patient 5 | Warthin Tumor | Parotidectomy | CN VII | S2e, S3 | 165 | Warthin Tumor |
| Patient 6 | Giant Cell Tumor | Parotidectomy, Mastoidectomy | CN VII | 1b, S2f | 120, 190 | Tenosynovial Giant Cell Tumor |

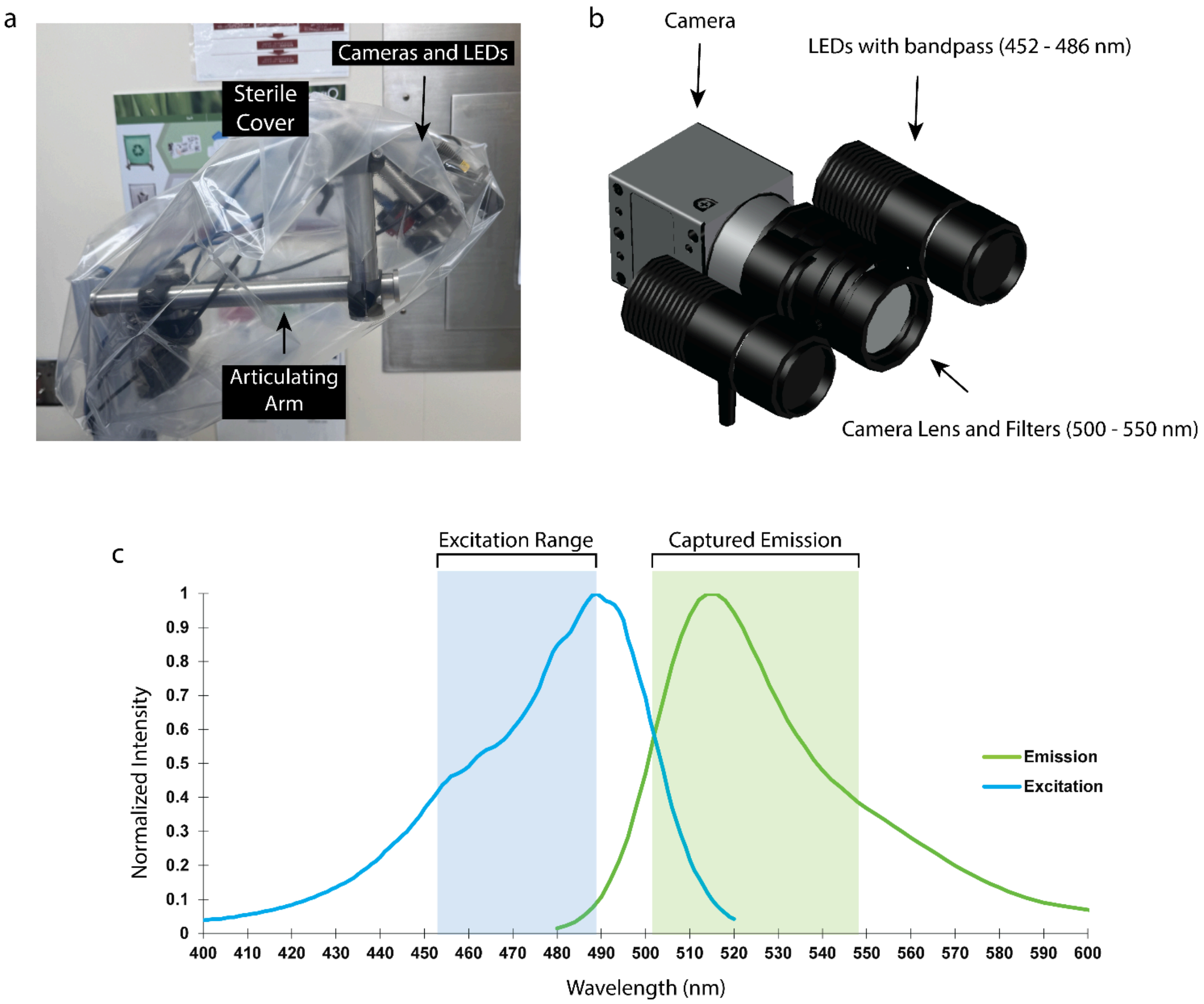


**Figure S1. Custom intraoperative fluorescence imaging system.** a) The custom imaging system consisted of a movable articulating arm, 2 LEDs for even illumination, and a 12 MP uncooled monochrome camera with a 35 mm lens. The entire system was covered with a drape to maintain sterility. b) 3D rendering of the imaging set up c) Excitation and emission spectrum of sodium fluorescein. Scales indicate the range of excitation from 452 to 486 nm and captured emission signal from 500 to 550 nm, determined by the full width at half-maximum of the filters used in the imaging system.

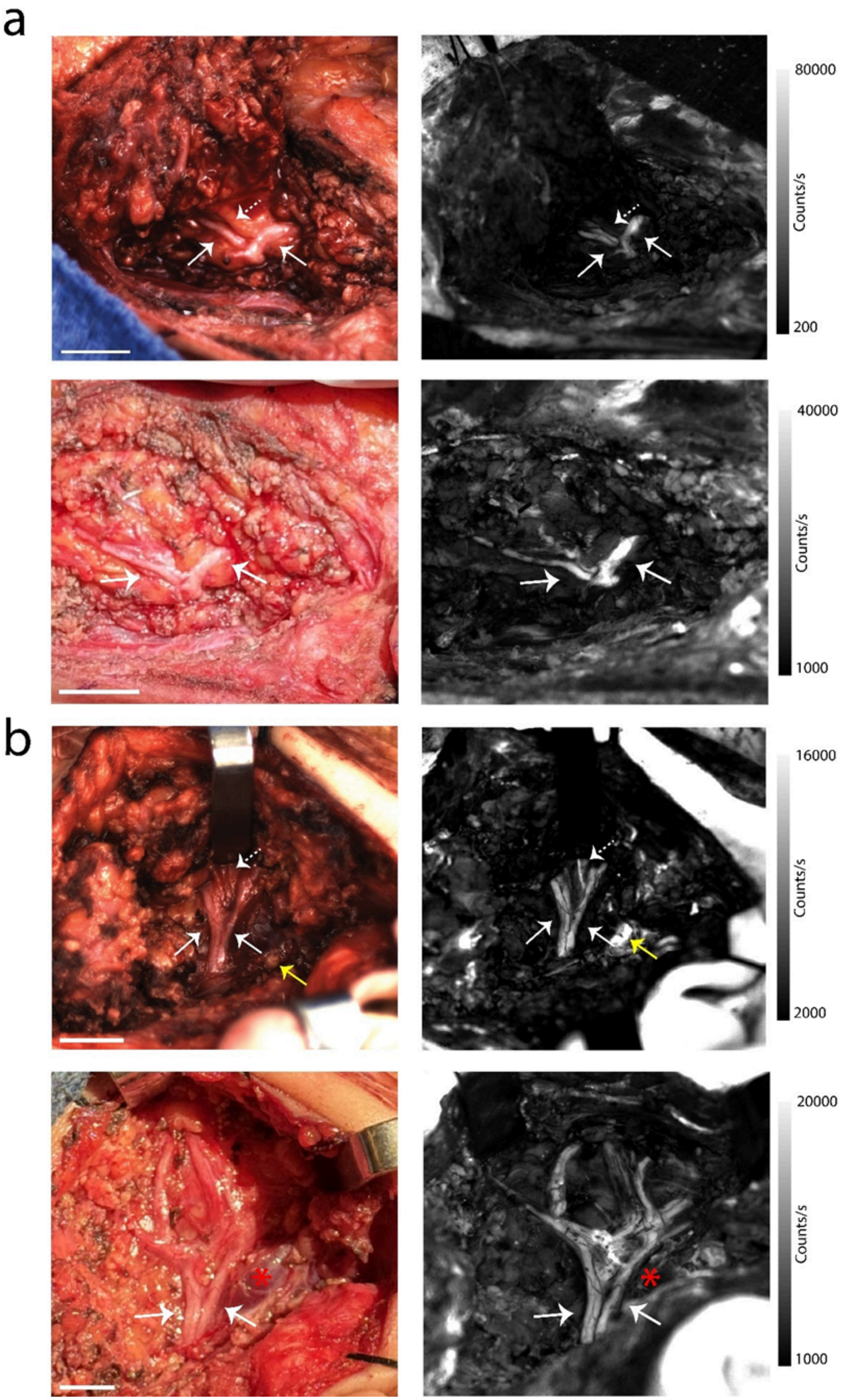

a
b
80000
200
40000
1000
16000
2000
20000
1000
Counts/s

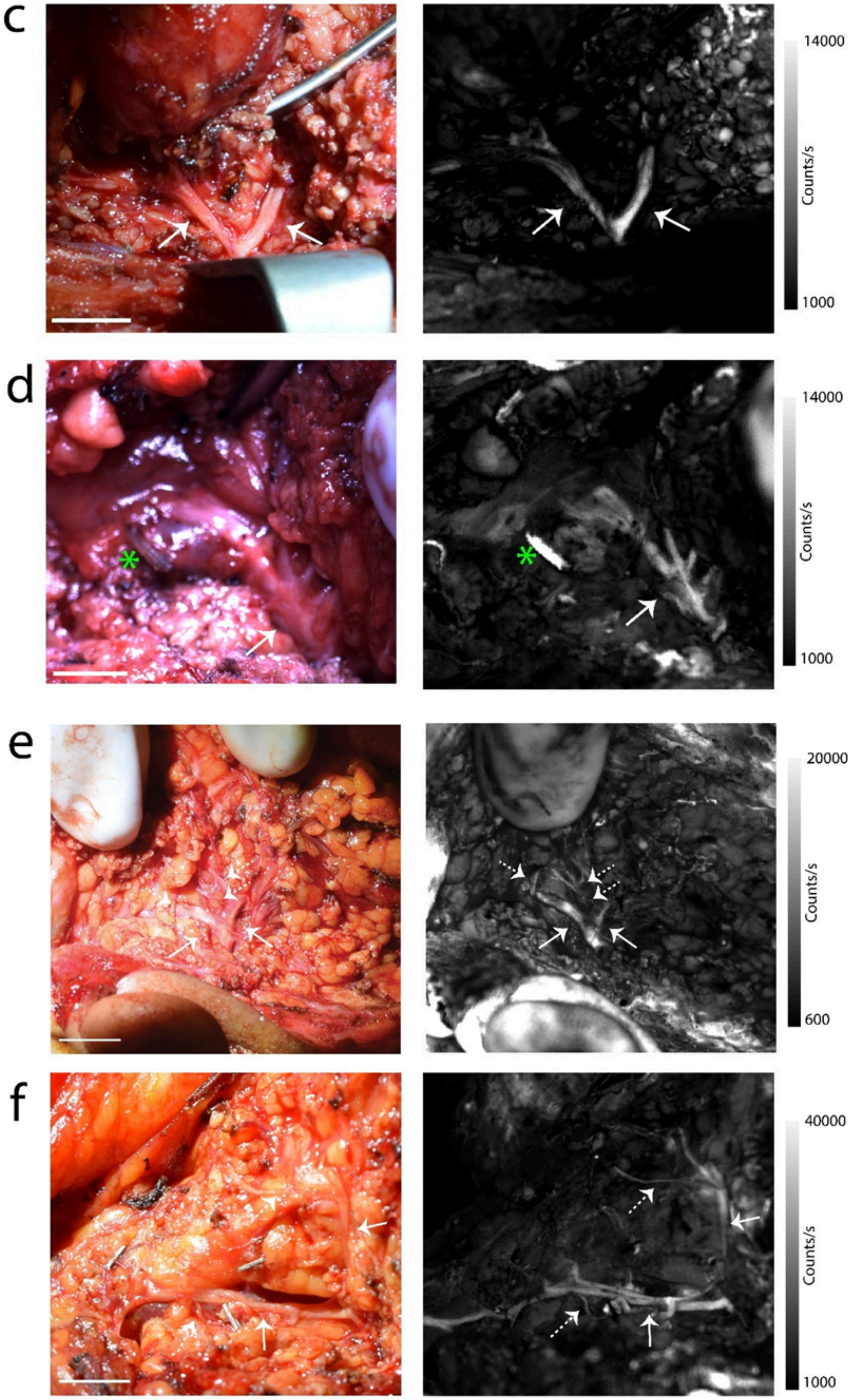
c
14000
Counts/s
1000
d
14000
Counts/s
1000
e
20000
Counts/s
600
f
40000
Counts/s
1000

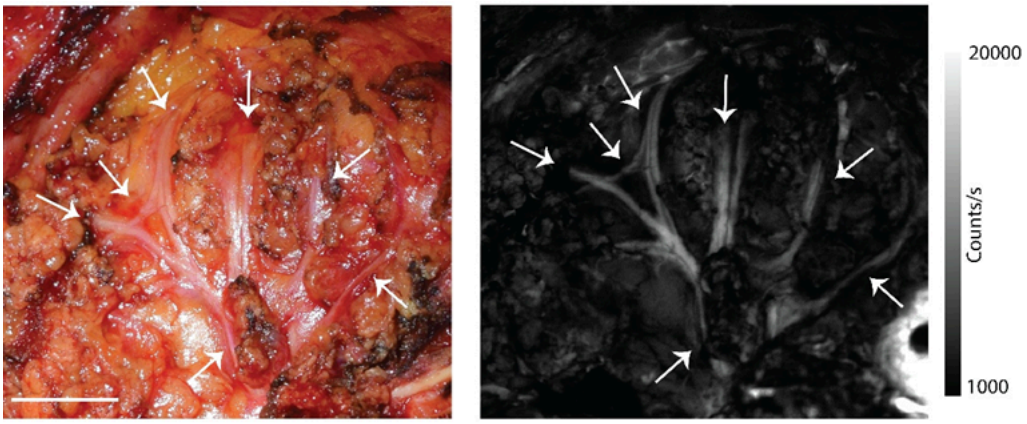


**Figure S2. Fluorescence nerve imaging of all patients enrolled in the Stanford clinical trial.** a-f) White light and corresponding fluorescence images of nerves. Each subfigure corresponds to a distinct patient. White arrows correspond to facial nerve, while dashed white arrows represent nerve branches with low contrast in the visible. Red stars correspond to retromandibular veins and green stars are surgical clips placed at end of retromandibular vein. Yellow arrow is lymph node. Scale bar 1 cm.

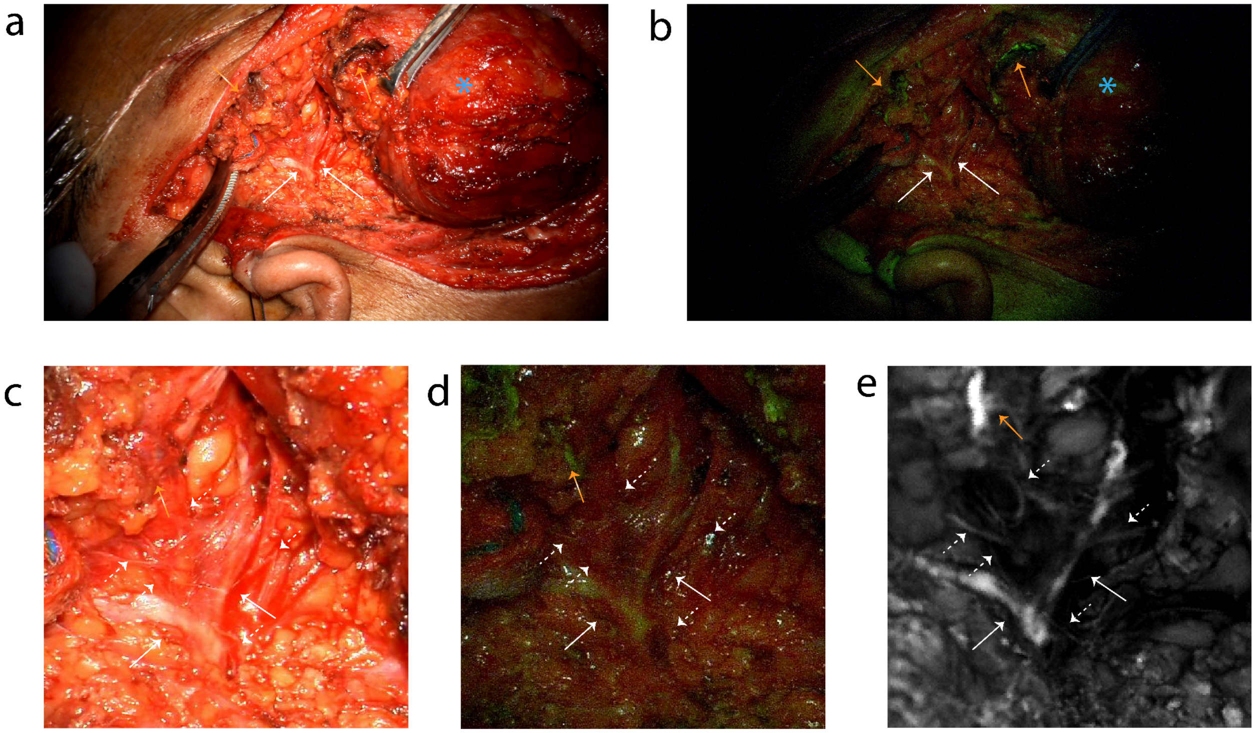


**Figure S3. Fluorescence nerve imaging with Zeiss Kinevo Yellow 560 system.** a) White light image b) Fluorescence image with Yellow 560 under auto setting c) Close up white light image of the exposed facial nerve d) Corresponding Yellow 560 image e) Fluorescence image with custom imaging system. White arrows correspond to the main branches of the facial nerve. White dotted arrows refer to small facial nerve branches. Orange arrows correspond to electrocautery cautery artifact. Blue stars are salivary gland tumor.

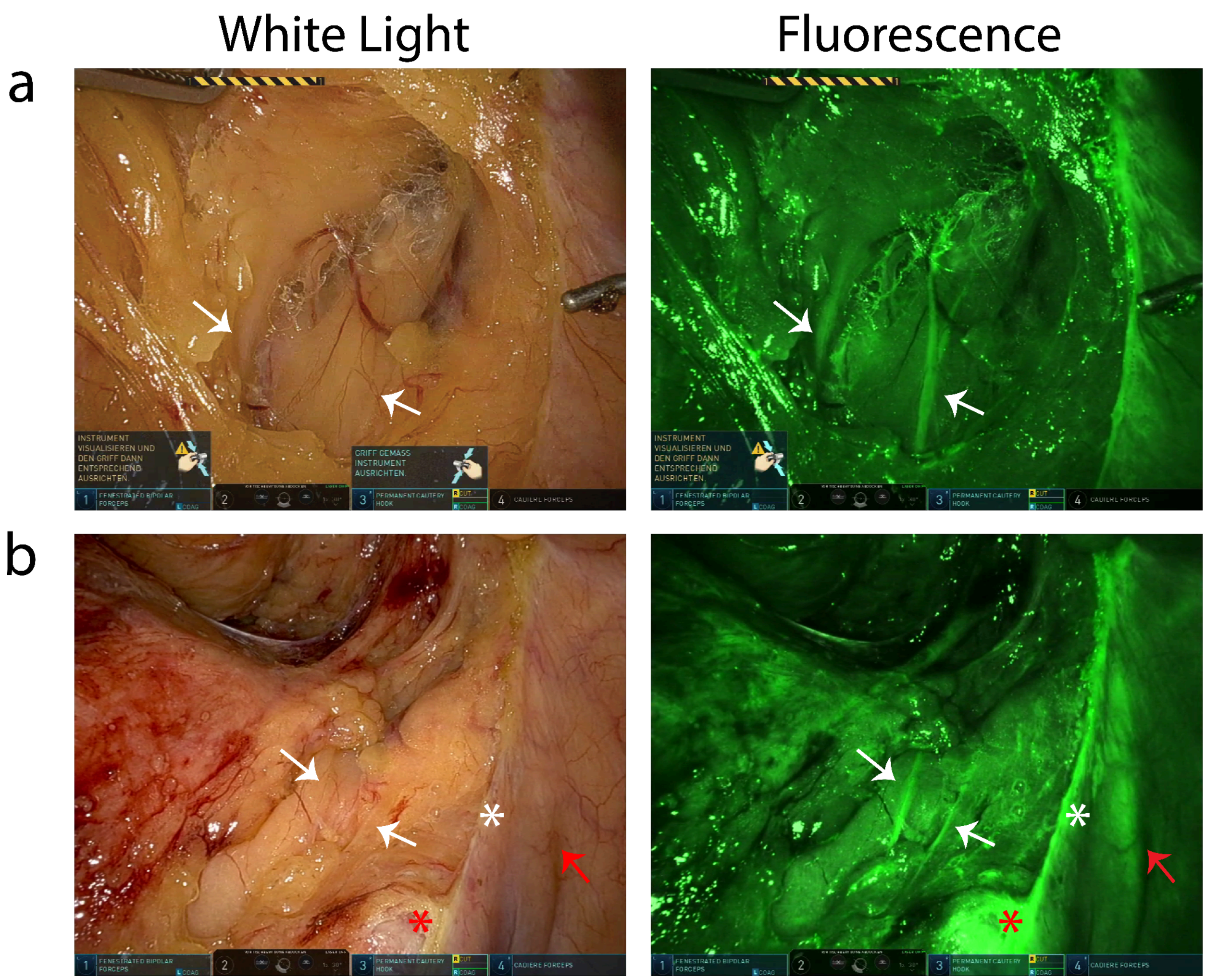


**Figure S4 Nerve imaging with Da Vinci Firefly endoscope.** a-b) Corresponding white light and fluorescence images with Da Vinci Firefly. White arrow corresponds to nerves of the superior hypogastric plexus. White star is margin of opened peritoneum in the small basin. Red stars represent iliaca dextra artery and red arrows represent ureter.

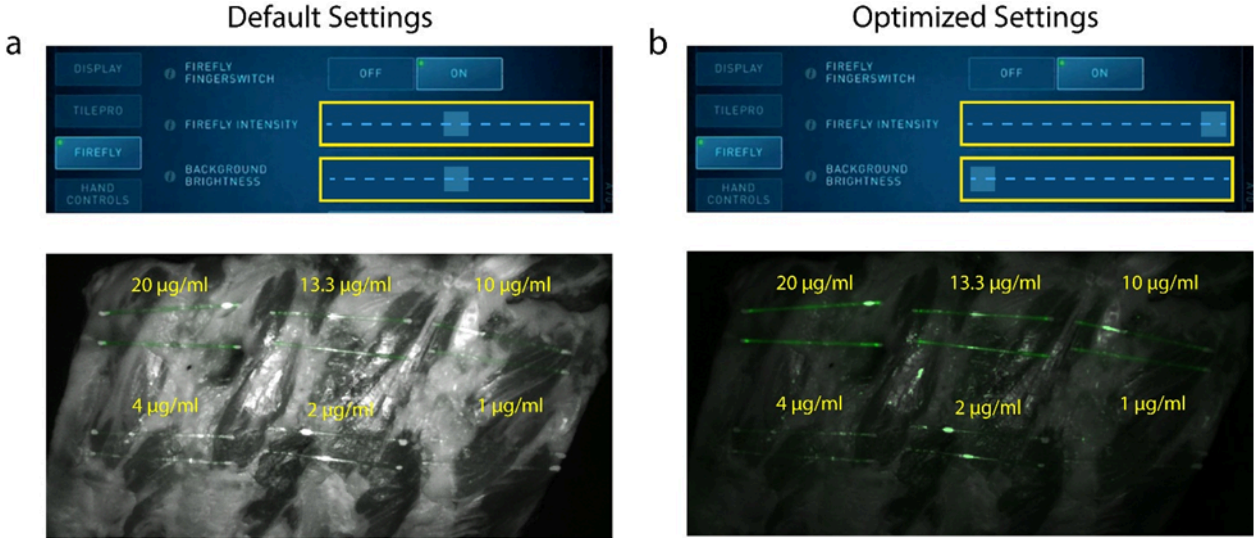


**Figure S5. Sensitivity comparison of the Da Vinci Firefly endoscope at default and optimized setting.** a) Fluorescence detection of sodium fluorescein dilutions in capillary tubes with default settings of the Da Vinci endoscope. These images are dominated by tissue reflections, which prevented the detection of concentrations of 4 μg/ml and lower. b) Optimization of the Da Vinci system settings – maximizing Firefly intensity and minimizing white light intensity (optimized settings) – reduced non-fluorescent light and improved detection of lower concentrations. Each capillary doublet represents a different concentration of fluorescein diluted in saline.

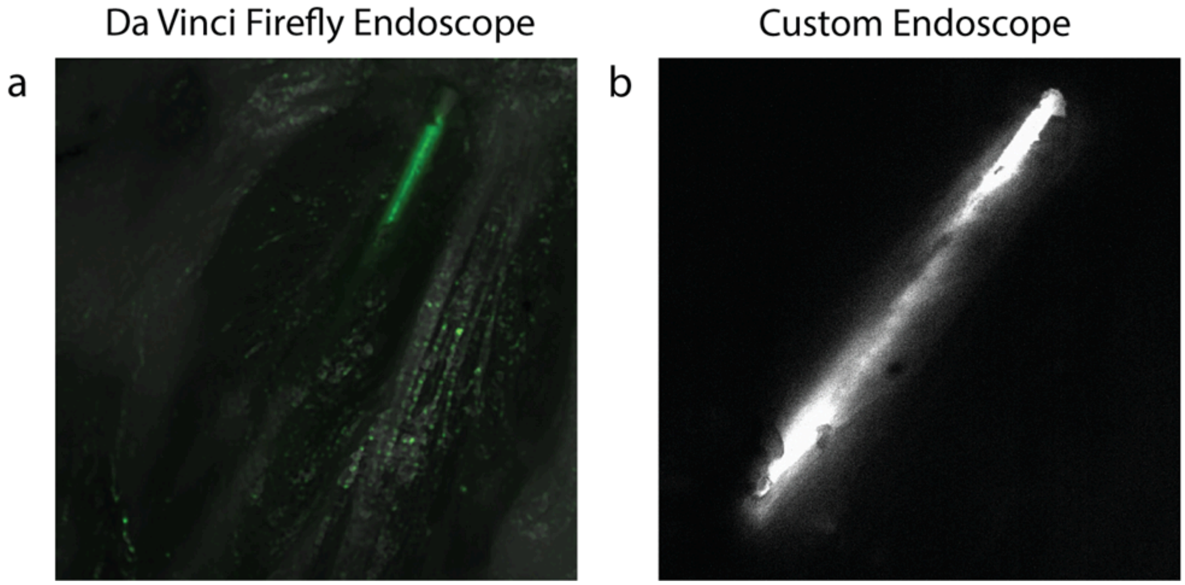


**Figure S6. Comparison of Da Vinci Firefly endoscope and custom endoscope for embedded sodium fluorescein capillary tube under 1 mm of tissue.** a) Fluorescence detection of capillary tube with endoscope using Da Vinci Firefly endoscope. b) Detection of capillary tube using custom endoscope.

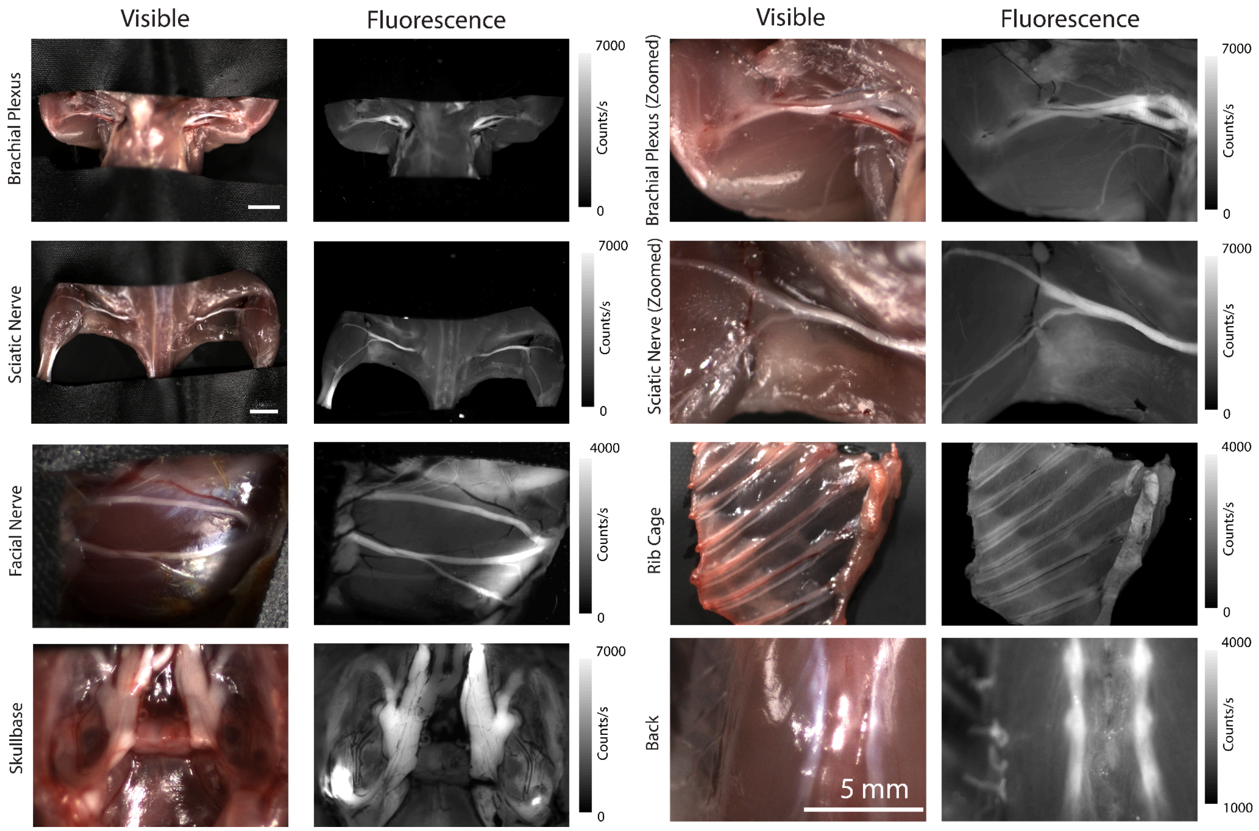


**Figure S7. Pan-body survey of nerves in murine models after administration of sodium fluorescein.** Pan-body survey of nerves comparing white light and fluorescence after fluorescein administration. Scale bar 5 mm.

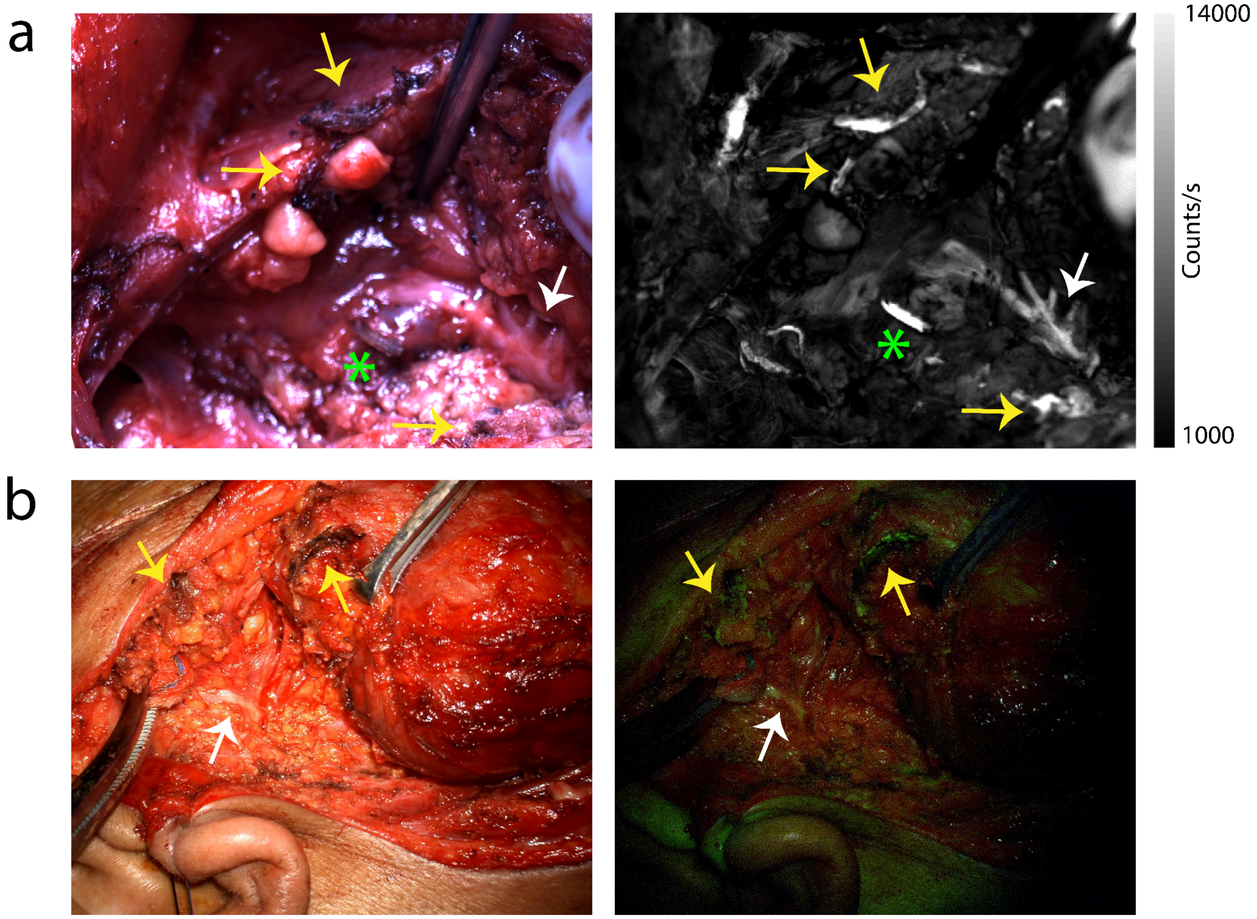


**Figure S8 Electrocautery artifacts highlighted with sodium fluorescein imaging.** a-b) Electrocautery artifacts (yellow arrows) were also detected when imaging nerves (white arrow) with sodium fluorescein imaging. Green star represents surgical clip.
